# Continuous monitoring of the use of molecular diagnostics in oncology: advancing towards a learning health system

**DOI:** 10.1101/2025.09.07.25335287

**Authors:** Kees C.W.J. Ebben, Femke J. C. Jacobs, Robin Pasman, Carin Louis - van den Broek, Cornelis D. de Kroon, Anthonie J. van der Wekken, Paul A. Seegers, Quirinus Voorham, Eline Heidenrijk, Jurrian van der Werf, Silvia van der Flier, Sahar Barjesteh van Waalwijk van Doorn - Khosrovani, Marjolijn J.L. Ligtenberg, Robby E. Kibbelaar, Wendy W.J. de Leng, Ed Schuuring

## Abstract

Molecular Diagnostics (MDx) is integral to the personalization of cancer treatment, offering both prognostic and predictive insights that inform therapeutic decision-making. However, the rapid advancement of MDx in oncology comes with significant challenges for the development, implementation, and adherence to clinical guidelines. A systematic approach is required to monitor clinical practice, including the utilization of MDx and the evaluation of patient outcomes. Such an approach can facilitate evidence-based updates to clinical guidelines, enhance adherence, and increase transparency in healthcare decision-making. This study aimed to develop a proof-of-principle practice monitor for MDx using two showcases, non-small cell lung cancer (NSCLC) and endometrial cancer (EC).

Adopting a socio-technical framework, stakeholders were engaged to ensure alignment of the monitor’s content with clinical needs. Critical clinical questions were identified, and computational decision algorithms were developed based on current guidelines and expert consensus. Nationwide real-world data registries were analyzed to assess the availability, quality, and completeness of data in relation to the clinical questions and decision algorithms.

Variables included in decision algorithms were largely present for NSCLC and EC, confirming the feasibility of nationwide monitoring. While some clinical questions could be fully or partially addressed with the available data, many could not. This was due to fragmented data, inconsistent coding practices, and the use of non-standardized free-text entries. Accordingly, this initiative addresses key challenges related to data standardization, computational representation of guidelines, and variability in clinical reporting. With strong national databases and infrastructure already in place, successful implementation now hinges on focused multidisciplinary and multi-organizational collaboration and a learning community to sustain ongoing progress. This approach assessed the feasibility and challenges of continuous evaluation of MDx practice in a proof-of-principle practice monitor, as an important step towards a Learning Health System.

## 1. Introduction

Molecular Diagnostics (MDx) plays a crucial role in personalizing cancer treatment by the identification of specific molecular characteristics (biomarkers) in tumors to guide the selection of (targeted) treatments and to provide prognostic information(1). The rapid development of new targeted therapies and growing number of predictive biomarkers and diagnostic tests underscore the growing importance of MDx in modern oncology(2).

However, the rapid pace of advancements in MDx has outstripped the ability of traditional clinical practice guidelines to remain actual (3), resulting in unequal access to MDx for patients. In the Netherlands, this challenge prompted the establishment of the committee of Clinical Essential Targets (in Dutch ‘commissie Klinisch Noodzakelijke Targets’, in short ‘cieKNT’), which reports for most tumor types the current availability of targeted drugs and immunotherapeutics and the (molecular) targets that are associated with these treatment options in KNT-lists (4). The KNT-lists describe predictive and prognostic biomarkers that should be tested to identify patients that benefit from currently available (systemic) targeted and immune-therapies. The aim of the cieKNT is to support healthcare providers (including physicians, pathologists, and clinical scientists in molecular pathology) and to ensure equitable access for all cancer patients in the Netherlands to currently available targeted therapies and appropriate predictive and prognostic biomarker testing. While guidelines rely on evidence-based medicine (EBM) methodologies, the KNT-lists are developed through a more consensus-driven process, making them complementary yet distinct tools. Practical application of these instruments should be examined to ensure their effectiveness in clinical settings (5).

Beyond the challenges of keeping these instruments up to date, prior studies have revealed suboptimal testing rates and regional variations in guideline adherence, indicating disparities in patient care in the Netherlands (6–12). These findings highlight the need for regular systematic evaluation of the application of MDx in daily clinical practice to ensure equal access and optimal care delivery. While existing studies provide valuable insights, their sporadic nature and reliance on outdated data limits their ability to drive continuous improvement. A national, systematic and technically integrated approach to continuous collecting and analyzing data on MDx usage, subsequent treatment strategies, and patient outcomes is thereby essential (13).

These issues exemplify that MDx is an area in need of a Learning Health System (LHS). An LHS is a transformative approach designed to continuously improve healthcare by integrating clinical practice, knowledge representations, and real-world data (RWD) (14). At its core, an LHS learns from each patient interaction, aligning science, informatics, incentives, individual patient outcome and culture to foster ongoing innovation and improvement (15). New knowledge is seamlessly embedded into the delivery of care, while the care process itself generates valuable insights. To fully realize the potential of a Learning Health System (LHS), coordinated collaboration across multiple organizations and disciplines—including all relevant stakeholders—is essential. This entails dismantling silos between key components such as clinical practice guidelines, source data capture (e.g., electronic health records), and the integration of real-world data from (national) registries. Notably, in the Netherlands, multiple national healthcare databases exist, however, data required for real-world monitoring is fragmented across multiple databases. By enabling open information flow and connectivity between these components—facilitated by open standards—LHSs are needed to drive continuous learning, implement relevant findings and improve patient outcomes (16).

This study aims to identify the barriers and facilitators for continuous monitoring of the use of MDx in oncological clinical practice while assessing its feasibility for seamless integration into an LHS. This involves a socio-technical design process and the development of technical components (17). The socio-technical track emphasizes engagement with diverse stakeholders to ensure that the practice monitor addresses the needs of all key users (18). The study identifies challenges related to data standardization and centralized data collection, knowledge representation (e.g. guidelines), and clinical integration through a hands-on approach, delivering a proof-of-principle to demonstrate feasibility in this rapidly evolving field of growing number of new targeted therapies and their associated new targets. By combining robust stakeholder engagement with technical innovation, this study seeks to create a sustainable direction for improving the application of MDx in oncology within an LHS framework.

## 2. Materials and Methods

### 2.1 Study Design

The *Practice Monitor for Molecular Diagnostics* project was initiated and funded by the Dutch National Health Care Institute to evaluate and improve MDx practices in oncology. The project used non-small cell lung cancer (NSCLC) and endometrial cancer (EC) as case studies to test the feasibility of the practice monitor. These cases served to explore how standardized MDx data could be integrated into a practice monitor as part of an LHS (see Figure 1), while identifying opportunities for refinement. The study was conducted between January and October 2024.

**Figure 1.**
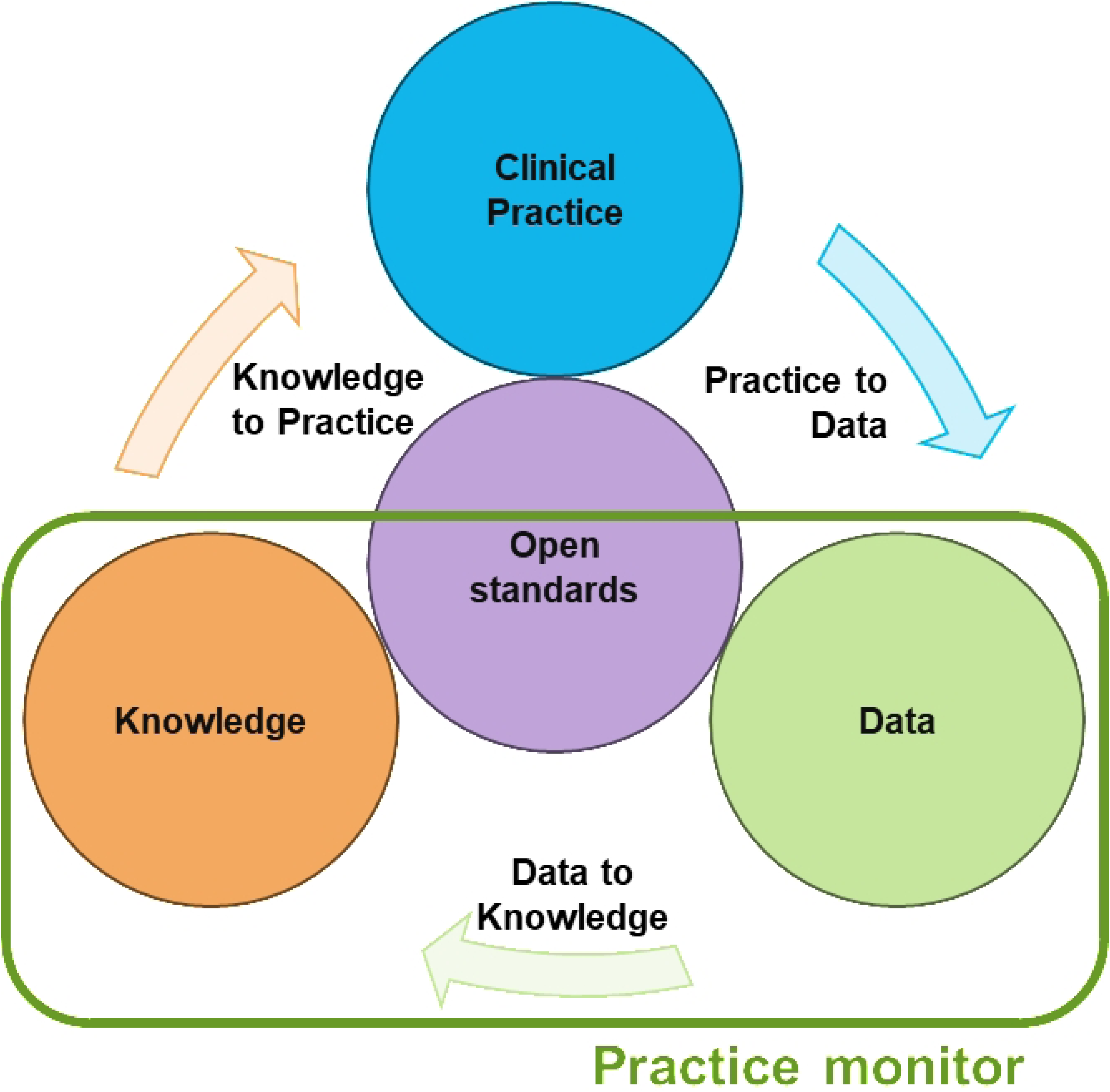
A Practice Monitor as Part of a Learning Health System. A Learning Health System consists of four key elements (circles) connected by processes (arrows), with the flow of information at its core. The practice monitor primarily focuses on transforming data (from existing nationwide registries) into knowledge.

### 2.2 Socio-Technical Approach

A socio-technical approach was adopted to ensure that the practice monitor addressed clinical, technical, and organizational needs (18). The project was guided by a multidisciplinary working group, including healthcare professionals, patient advocates, representatives from data and knowledge parties, policy organizations, professional associations, guidelines committees, research institutions, and an HTA expert (see Table 1 and Figure 2.).

**Figure 2.**
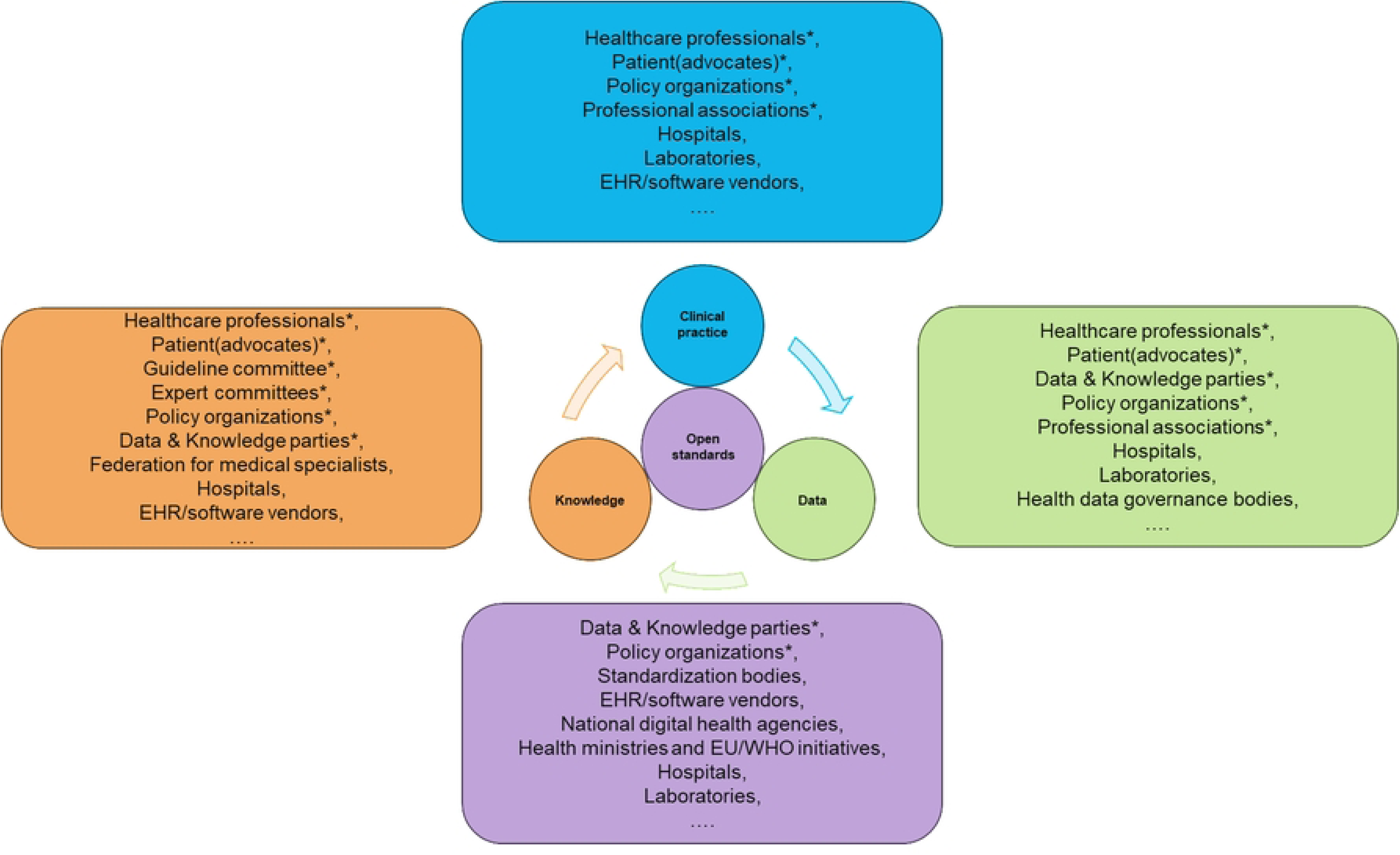
A Learning Health System (LHS) continuously improves healthcare by integrating clinical practice, data, knowledge, and open standards. Each component relies on input, contributions and usage from a diversity of actors. This figure provides an (non-exhaustive) overview of stakeholders within a nationwide LHS, with representatives marked with an asterisk having participated in this project. EHR = Electronic Health Records, EU = European Union, WHO = World Health Organization, IT = Information Technology

**Table 1.** Overview of participating organizations and stakeholder categories involved in the Practice Monitor Molecular Diagnostics project. The table categorizes contributors by their primary domain, including patient organizations, medical specialties, national committees, data and policy institutions, and academic centers.

| Category / Disciplines | Organizations |
| --- | --- |
| **Patient Advocates** | - Dutch Federation of Cancer Patient Organizations (NFK)<br>- Olijf Foundation<br>- Lung Cancer Netherlands Foundation |
| **Professional Associations / Healthcare professionals** | - Dutch Pathology Society (NVVP)<br>- Clinical Molecular Biologists in Pathology<br>- Dutch Association of Physicians for Pulmonary Diseases and Tuberculosis (NVALT)<br>- Dutch Society for Obstetrics and Gynaecology (NVOG) |
| **Expert Committees** | - Committee for Clinical Essential Targets (cieKNT)<br>- Committee for the Assessment of Oncological Diagnostics (cieBOD)<br>- Committee for the Assessment of Add-on Medicines (cieBAG)<br>- Guideline committee Endometrial Cancer<br>- Guideline committee Non-Small Cell Lung Cancer |
| **Data and Knowledge parties** | - Palga Foundation (Pathological Anatomical National Automated Archive)<br>- Dutch Institute for Clinical Auditing (DICA)<br>- Netherlands Comprehensive Cancer Organization (IKNL) |
| **Policy Organizations** | - Dutch Association of Health Insurers (ZN)<br>- National Health Care Institute (Zorginstituut Nederland) |
| **Guideline experts, HTA Experts, and Research Institutions** | - University Medical Center Groningen (UMCG)<br>- Leiden University Medical Center (LUMC)<br>- Amsterdam University Medical Centers (Amsterdam UMC)<br>- Radboud University Medical Center (RadboudUMC) |

To structure the development process, three key meetings were held.

- In the initial meeting, project goals were defined, and relevant clinical questions were identified.
- A subsequent progress meeting focused on governance frameworks for an MDx-based practice monitor, prioritized clinical questions, and outlined requirements for applicable technical components.
- The final meeting refined the requirements and explored future implementation strategies.

### 2.3 Formulating Clinical Questions

A crucial step in developing the practice monitor was formulating relevant clinical questions to guide the analysis of real-world data. This process mirrored the formulation of research questions in guideline development but was specifically tailored to leverage real-world data for ongoing knowledge generation. Instead of defining questions to establish new evidence through clinical studies, the focus was on extracting insights from yet existing data. To ensure clinical relevance, the working group was asked to collaboratively reflect on proposed key areas where data-driven insights could support decision-making.

### 2.4 Development of Technical Components

The technical development of the practice monitor was structured around computational knowledge models (structured, algorithm-based protocols) for MDx, standardized clinical reporting templates, and RWD analyses. The development of technical components was structured into three distinct work packages.

#### 2.4.1 Clinical decision algorithms

The first technical component involved the transformation of national clinical practice guidelines (authorized by the Scientific Societies of the Federation of Medical Specialists) and the supplementary KNT-lists for NSCLC (stage IV) and EC (all stages) with a relation to MDx into computer-interpretable Clinical Decision Algorithms (CDAs). This process required identifying key clinical variables, specifically those defined in the guidelines and KNT-lists: (1) patient and disease characteristics along with their values, (2) interventions, and (3) healthcare outcomes. These variables were assigned with codes according to the Systematized Nomenclature of Medicine Clinical Terms (SNOMED CT) terminology system to standardize and enhance interoperability and develop a semantic information standard (19). CDAs consist of a top node that defines the population of interest and specific point in the care pathway. This is followed by intermediate nodes representing patient and disease characteristics, which are interconnected by connectors, each corresponding to a specific value of the characteristic. These pathways ultimately lead to leaf nodes that contain the recommended clinical actions for the defined subpopulations (20). To ensure robust knowledge representation, multiple dedicated work sessions were conducted with subject matter experts, patient representatives, and informaticians.

#### 2.4.2 Standardized, structured clinical reporting

To advance an LHS, structured templates were developed for use in multidisciplinary team (MDT) discussions, ensuring consistency between clinical reporting and knowledge representation. These templates were aligned with existing information standards to improve data capture and support decision-making in oncological care. The development of a standardized, structured report (SSR) starts with a comprehensive analysis of the care process, and the specific information needs for the given decision-making moment. Next, for optimal configuration of clinical reporting, a structure of the template is first established in the form of grouping various concepts. Finally, the concepts, as recorded in a semantic information standard, are incorporated, and conditionality between concepts is introduced (21). During the key meetings the relevance and content of these forms were discussed with the working group. Implementation of these forms was not part of the project.

#### 2.4.3 Data accessibility and usability

The availability of nationwide datasets was assessed to determine their legal, technical, and functional accessibility. The included organizations maintain high-quality, nationwide health data registries that are essential for clinical research and healthcare evaluation in the Netherlands. The Palga Foundation provides a unique, population-wide pathology database with standardized reports from all Dutch pathology labs. The Dutch Institute for Clinical Auditing (DICA) oversees detailed clinical quality registries aimed at benchmarking and improving care. The Netherlands Cancer Registry (NCR), maintained by the Netherlands Comprehensive Cancer Organization (IKNL), offers national cancer incidence, treatment, and survival data. Also, usability analyses were performed to evaluate the extent to which these sources could contribute to the practice monitor. If data was accessible during the project period, it was obtained and analyzed for completeness and quality. Data quality is evaluated based on the level of detail, data types (structured vs. free-text), and its reusability for secondary purposes, ensuring its suitability for integration into the practice monitor. Collaboration with data experts from participating organizations ensured that data availability and usability were thoroughly assessed. Data from the NCR and Palga Foundation were accessed for research purposes on 30 July 2024. All data were provided in pseudonymized form, and authors did not have access to any directly identifying information of individual participants.

## 3. Results

### 3.1 Socio-Technical Design and Clinical Questions

Stakeholder engagement in the socio-technical design process highlighted the importance of monitoring which biomarkers are actually being tested in clinical practice and how this aligns with national recommendations. Based on these priorities, a set of key clinical questions was defined, forming the foundation of the practice monitor’s analytical framework. These questions focus on the use of molecular diagnostics (MDx), their influence on therapeutic decision-making, and observed patient outcomes. Table 2 provides an overview of these questions.

**Table 2.** The clinical questions identified by the working group as relevant to oncological molecular diagnostics.

| No. | Clinical Question |
| --- | --- |
| 1 | On which patients has molecular diagnostics been performed? |
| 2 | What type(s) of test(s) were used to detect molecular abnormalities? |
| 3 | Which biomarkers were tested? |
| 4 | Which clinically relevant biomarkers were identified? |
| 5 | Are patients receiving a treatment that aligns with the identified biomarkers? |
| 6 | What are the patients outcomes? |

### 3.2 Technical Components

We developed guideline- and KNT-list based Clinical Decision Algorithms (CDAs) for stage IV NSCLC and all-stage EC, revealing key clinical variables essential for decision-making. These include patient and disease characteristics, recommended interventions, and outcomes. The NSCLC CDA included 11 patient and disease characteristics (e.g., PD-L1 expression), 22 interventions (e.g., pembrolizumab), and one outcome -Overall Survival (see Supplement 1). The EC CDA included 19 patient and disease characteristics (e.g., age), 27 interventions (e.g., Hysterectomy), and likewise focused on overall survival as outcome (see Supplement 2).

Concept reporting templates for both NSCLC and EC were developed in alignment with the established information standard. These templates were designed to integrate seamlessly into the clinical workflow, and include all variables present in the CDAs. The information standards specify all relevant variables in this project, including their definitions, values or units, and, where available, corresponding SNOMED CT codes.

Dataset definitions from the Palga databank, as well as from the DICA and IKNL registries, were analyzed, revealing variation by cancer type and data category (see Table 3). For EC, patient and disease characteristics (19/19 datasets) and survival outcomes (1/1) were consistently reported, whereas intervention data showed partial coverage (20/27). In NSCLC, reporting was similarly complete for characteristics (11/11), interventions (22/22), and outcomes (1/1). However, substantial variation in reporting formats was observed across registries. A key distinction emerged between question-driven reporting (e.g., *“Is the patient over 60?”*) and data-driven reporting (e.g., *age: 63 years*). The latter enables broader reuse, as structured data elements can support multiple clinical questions and secondary purposes. Furthermore, no single source captured the full spectrum of clinically relevant data. Therefore, integrating multiple registries was essential to comprehensively address the formulated clinical questions.

**Table 3.**
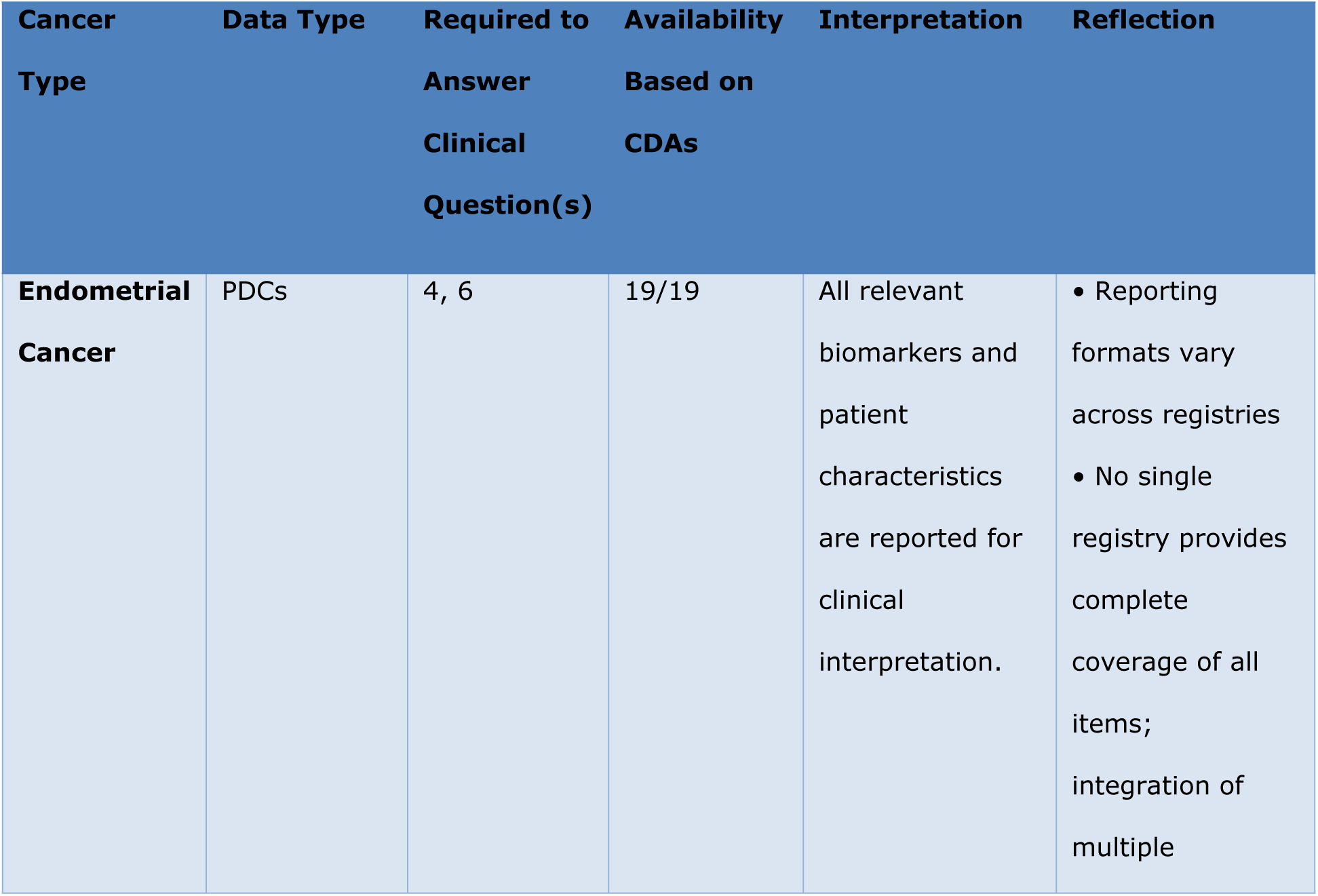
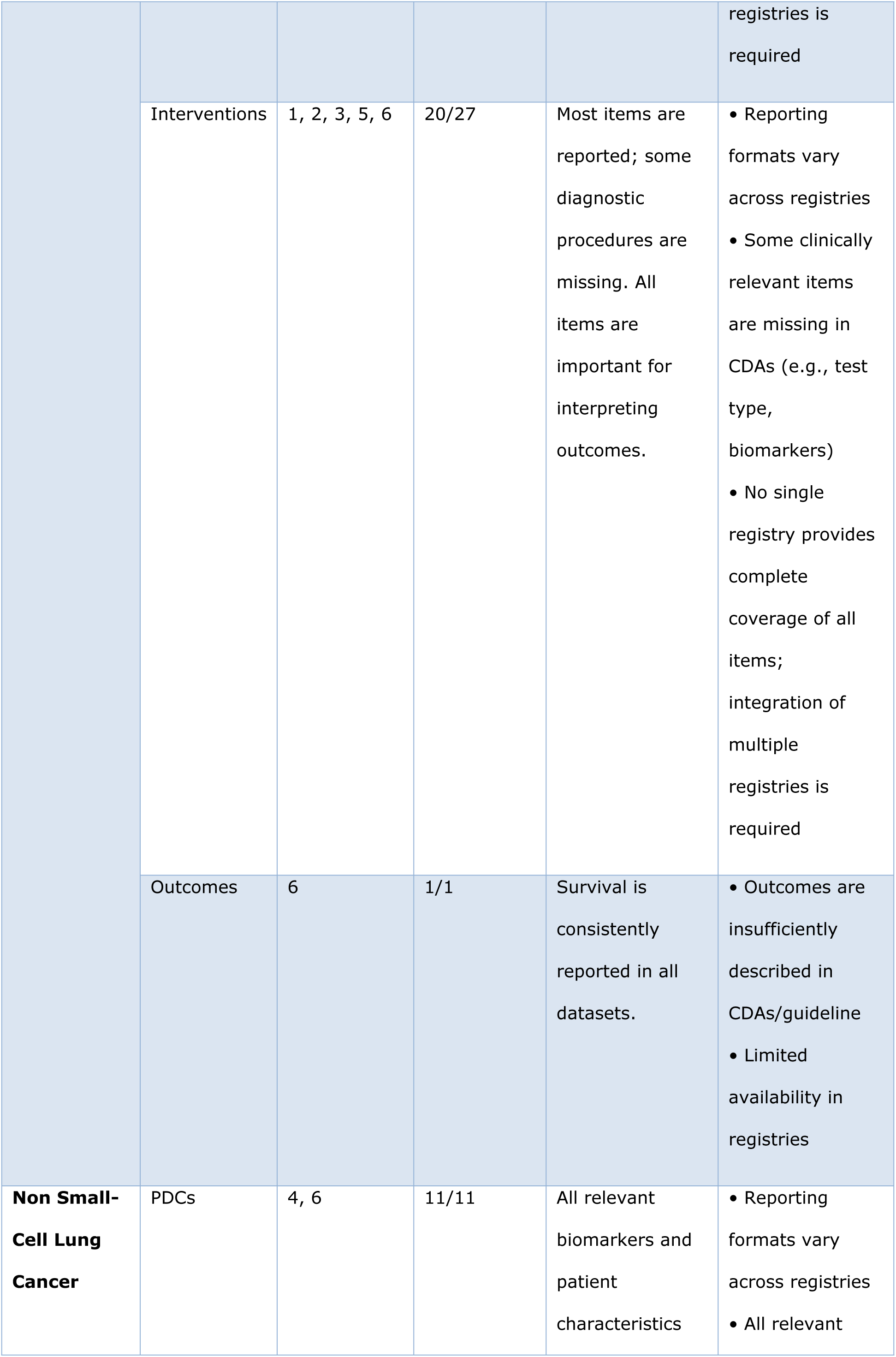
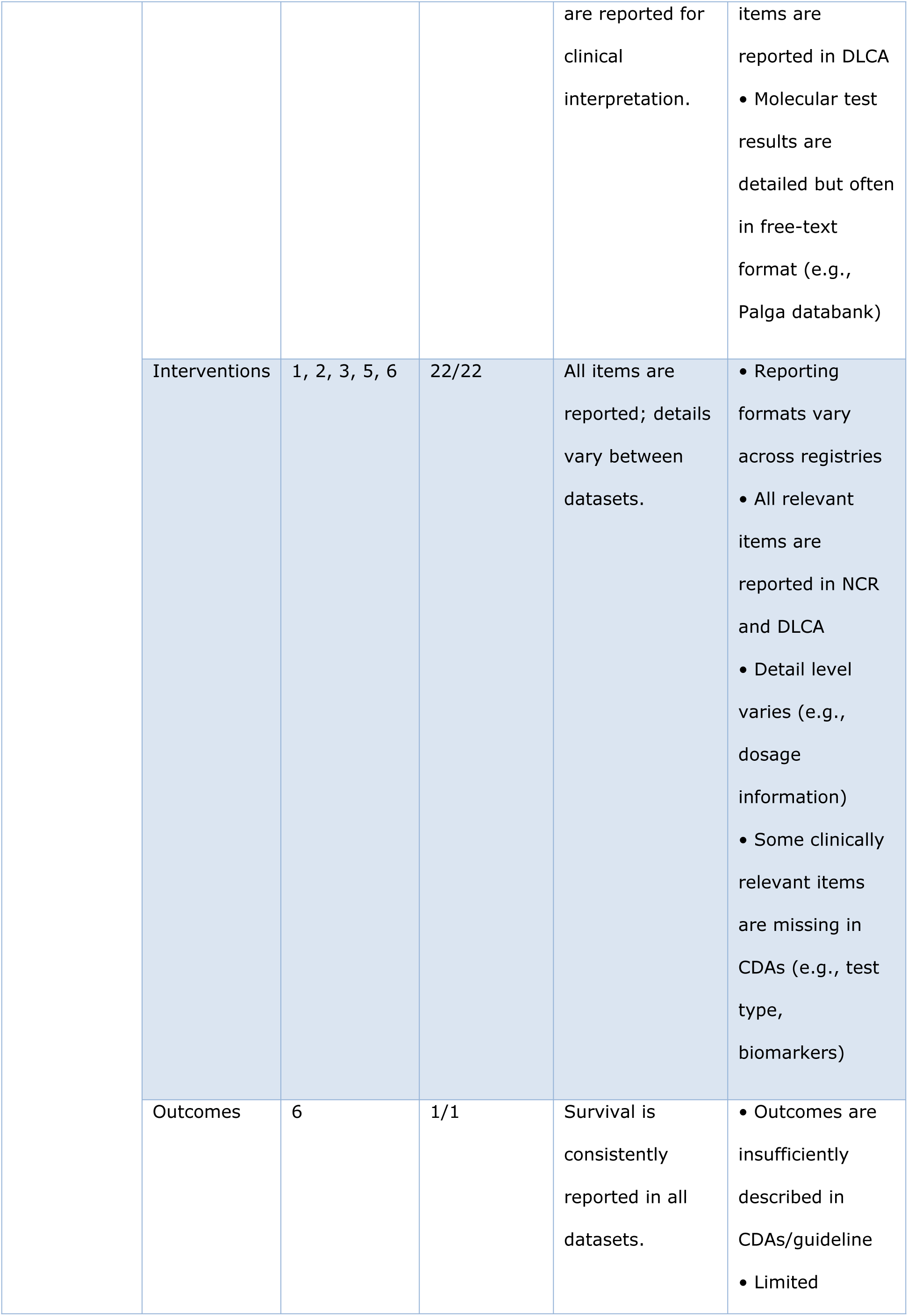
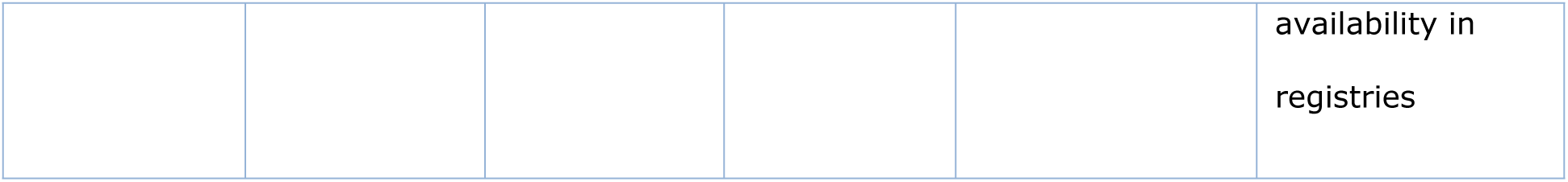
Interpretation and reflection on the data availability for the CDAs, and the usefulness for the clinical questions by cancer type and data type. CDAs = clinical decision algorithms; PDCs = Patient and Disease Characteristics; NCR = Netherlands Cancer Registry; DCLA = Dutch Lung Cancer Audit

Registry integration is at this point not possible for all registers inventoried. Furthermore, legal constraints prevented access to DICA data. These organizations require separate agreements with each participating hospital for data linkage, a process that proved infeasible within the project’s timeframe. Subsequently, an analysis was conducted on the available data, which was limited to the NCR and Palga datasets. In total, 12,489 patients diagnosed with NSCLC and 3,865 patients with EC in 2022 or 2023 were identified in the NCR. These records were subsequently linked to Palga data using pseudonyms based on patient identifiable data. However, data processing was challenged by inconsistent terminology (e.g. KRAS mutation, Lung KRAS G12C, KRAS), detail level of data (e.g. immunotherapy vs. pembrolizumab) and prevalent use of free-text entries (e.g. “…An activating mutation in KRAS exon 2 has been identified in the tissue…”).

Of the six aforementioned clinical questions (see Table 2), only one —extraction of demonstrated clinically relevant biomarkers— was fully addressed using the available data (see Table 3). In contrast, questions on test types and the specific biomarkers tested could not be addressed at scale. Questions concerning the alignment of treatment with identified biomarkers, the identification of patients undergoing molecular diagnostics, and patient outcomes were only partially addressable. In addition to the prementioned issues of inconsistent reporting, the use of free text and the challenges of data integration between registries, other encountered issues were the inconsistent use of standardized, structured reporting protocol in hospitals, and limited availability and quality of follow-up data.

## 4. Discussion

### 4.1 Importance of Findings

In the Netherlands, clinical data on molecular diagnostics (MDx) in oncology are recorded in independent, specialized registries, such as the PALGA databank for pathology data, the NCR for clinical cancer data, and DICA for clinical quality registries. Optimal monitoring of MDx application requires the alignment and integration of data from these three registries to create a comprehensive, interconnected dataset. This study demonstrates the technical feasibility of developing a practice monitor that evaluates the use of MDx in oncology, with the potential to support evidence-based guideline development, enhance policy-making for improved access, and advance the quality, efficiency, and transparency of cancer care. By combining real-world data (RWD) with clinical decision algorithms, the monitor provides a structured framework to inform clinical practice through real-world evidence. However, while this proof-of-principle confirms the technical viability of such a system, substantial challenges remain that must be addressed before valid, large-scale, and continuous monitoring can be realized.

Most importantly, developing a fully functional practice monitor will require a multi-organizational, multidisciplinary learning community. As experienced in this project, collaboration among stakeholders from clinical practice (healthcare professionals and patients), data experts, and guideline specialists is crucial to identify essential data, knowledge needs, and the interpretation and implementation of findings. In parallel, the availability and usability of real-world data must be enhanced through improved completeness, standardized and structured reporting, and interoperability across registries. Additionally, the integration of computational knowledge—including up-to-date clinical practice guidelines—into the system is crucial to enable automated and meaningful analyses. These key actions were identified in this study and have been captured in a jointly developed roadmap created with stakeholder input. To improve transparency and foster collaboration, both the publicly available roadmap and this paper propose similar recommendations. The roadmap, available via the National Health Care Institutes website (in Dutch), offers a practical guide for developing LHSs(21). Another deliverable of this project was the integration of RWD and computational guidelines into a prototype dashboard for evaluation purposes, which is separately detailed(22).

### 4.2 Study Limitations

There are several limitations of this proof-of-principle practice study. First, the prototype was constrained by the availability and quality of existing datasets, such as those from the NCR and Palga. Not all hospitals utilize standardized, structured pathological reporting templates, and the quality of these protocols needs improvement to enable data reuse. Besides, due to legal restrictions and the limited duration of this project, only limited data sources could be accessed, most probably affecting the comprehensiveness of the analysis. Moreover, this study relied on existing procedures, along with their associated limitations, for requesting data from the NCR and PALGA. However, implementing a sustainable monitoring system will require the establishment of a dedicated process, supported by appropriate legal and practical frameworks. Additionally, standardizing outcome data proved labor-intensive, requiring significant manual effort to extract and structure information from disparate, unstructured systems, highlighting a critical barrier to routine and scalable implementation. Moreover, the inherent time required for implementing interventions, collecting RWD, and processing the data for analysis introduces a latency into the system, which is a common challenge in real-world monitoring systems. Nevertheless, using a practice monitor to obtain real-world insights provides potential to reduce delays compared to the often-cited research-to-practice gap, that is inherent to research in a clinical trial setting (23). This shorter cycle time is crucial for improving equity and clinical outcomes more swiftly (24).

### 4.3 Future Directions

A cornerstone of any successful LHS is the establishment of a learning community(25), particularly for health topics like MDx in oncology. This community unites diverse stakeholders around shared goals to address challenges such as rapidly advancing diagnostic technologies and data integration. By fostering collaboration, learning communities ensure that clinical workflows are guided by actually available evidence while generating insights through continuous evaluation of real-world data. In this project, stakeholder-defined clinical questions shaped the design of a practice monitor for MDx, reflecting real-world needs. This learning community can play a key role in determining which (level of) information should be collected for practice monitoring with appropriate governance and sustainable finances. Regular engagement facilitated the identification of barriers, the development of actionable solutions, and the iterative refinement of tools such as dashboards, computational decision algorithms, and structured clinical reporting templates. Learning communities play a crucial role throughout the entire practice monitor cycle, from discovery to implementation, fostering scalability, promoting standardization, and ensuring shared accountability(25). Figure 2, developed based on the experiences from this project and existing scientific literature (17, 26–28), presents an overview of the key actors involved in a nationwide LHS. Notably, this study did not include experts in artificial intelligence, or large language models, stakeholders whose involvement could strengthen future monitoring efforts. The optimal composition of a learning community varies by use-case.

A key next step is the harmonization of clinical documentation with guideline-based data elements, ensuring a consistent flow of information across the care continuum (29). Standardized, structured clinical reporting templates and information standards can support interoperability and facilitate seamless integration into clinical workflows (30). To fully realize the value of clinical data, it is important to distinguish between its use for direct patient care, for (fundamental) research (e.g. investigating novel biomarkers), and for healthcare monitoring and evaluation, each with distinct requirements and implications for data governance. This project specifically focused on healthcare monitoring and evaluation, specifically requiring the long-term usability of data and a well-established data-infrastructure to support this. Such initiatives fall within the domain of the informatics community, highlighting the crucial role of this discipline in articulating the relevance of an LHS (27). Additionally, strengthening governance structures is crucial to ensure clear ownership, responsibility, and transparent agreements on data access. Establishing a dedicated governance framework for the practice monitor within existing healthcare governance structures will safeguard data integrity while promoting broader adoption.

A functional practice monitor requires high-frequency updates of clinical data, which can only be achieved through a well-designed and interoperable data infrastructure. Such an infrastructure must enable real-time or short-cyclic data exchange, ensure semantic consistency across sources, and support automated data capture from routine clinical workflows. To ensure scalability and sustainability, investment (e.g. financial, human and technical) in this kind of infrastructure is essential. Key challenges—inconsistent terminology, and the absence of standardized protocols—must be addressed. Although the national PALGA protocols for molecular diagnostics were primarily designed for clinical documentation, multiple studies have already demonstrated their reusability for research and monitoring (6, 9–12, 31). The current limitation lies in the unstructured nature of reporting, which makes linkage to other databases labor-intensive. Ongoing initiatives, such as the phased implementation of a pathology protocol module in several laboratories, are expected to enable more structured reporting and thereby facilitate broader and more efficient reuse. Limitations were in line with earlier findings(8, 32), and include inconsistent use of standardized, structured reporting across hospitals, lack of standardization in data formats and terminology, incomplete or missing fields, reliance on unstructured narrative text, and limited contextual metadata required for accurate interpretation. Enhancing data integration—across all sources—via national linkage mechanisms and open standards can enable more comprehensive and timely monitoring. Additionally, accelerating the standardization and technological integration of electronic health record systems will reduce manual effort and significantly improve data availability (33). Also, a major advancement lies in the creation of guideline based decision support systems integrated within clinical workflows, which can facilitate the translation of knowledge into practice (34).

Adherence measurement can be approached at multiple levels, ranging from transparency and simple reporting to active monitoring and quality assurance. Transparency involves making data on guideline compliance visible to stakeholders, while monitoring enables ongoing tracking of adherence trends over time. More advanced levels include systematic quality checks and feedback loops that support targeted improvements. Recognizing and implementing these layers is crucial for a practice monitor to not only assess compliance but also drive meaningful enhancements in clinical care.

### 4.4 Transition to a Learning Health System

The practice monitor represents a critical step towards the development of a fully integrated LHS where RWD and computational guidelines create a dynamic ecosystem of continuous knowledge generation and application. By leveraging federated platforms (which enable collaborative model training across institutions without sharing raw patient data, thereby allowing for privacy-preserving analyses) the practice monitor can facilitate closed-loop feedback between clinical practice and knowledge representation. This system not only enables the seamless exchange of data for aggregated analyses but also emphasizes the iterative refinement of knowledge representations based on real-world evidence.

Success in transitioning to an LHS requires coordination of semantics, including the definition of concepts within computational guidelines and the development of standards (35, 36). Standardized, structured clinical reporting is essential to ensure that collected data can be easily integrated and analyzed (37). High-quality registration enhances data reusability, both for direct clinical applications and for secondary purposes such as (international) research, quality improvement, and policy-making. In this context, the use of templates for standardized, structured clinical reporting is a crucial step in evolving from a practice monitor to a fully functional LHS. Additionally, the implementation of decision support tools at the point of care will be essential for translating research into practical improvements in patient outcomes. The integration of RWD directly from clinical practice, combined with scientific evidence from computationally-driven guidelines, creates a continuously evolving system that adapts to new knowledge and supports optimal patient care(38). This adaptive approach positions the practice monitor MDx as an essential basis for evaluating and refining current clinical practice, supporting learning from what is already implemented. While it does not yet enable the discovery of novel insights, it lays the foundation for a future in which structured, high-quality molecular data can be leveraged to drive deeper learning and innovation in oncology care and beyond.

## 5. Conclusion

This study establishes a foundation for monitoring MDx use in oncology in the Netherlands by integrating real-world data with computational clinical decision algorithms to systematically assess guideline adherence and patient outcomes. To fully accomplish the practice monitor’s potential for advancing oncology care, key challenges related to data-availability and -usability must be addressed.

Successful implementation requires strengthening data infrastructure and fostering multi-organizational collaboration among clinicians, clinical laboratory scientists, data analysts, informaticians, policymakers, health insurers and patients. Embedding the practice monitor within a learning health system will enable continuous insights from MDx applications, while building a multidisciplinary learning community is essential to drive ongoing improvement in optimized oncology care and patient outcome.

## Data Availability

Data cannot be shared publicly because of privacy and data protection regulations. Data are available from IKNL and PALGA through their standard procedures for data requests, for researchers who meet the criteria for access to confidential data. Requests can be made via the respective institutional access processes, referring to IKNL request number 24-00191 and PALGA request number 2024-64.

## Acknowledgements

The authors would like to sincerely thank Lotte Hermsen, Bert van Nistelrooij, Ilse Verstijnen, and Yoka Kusumanto from the National Health Care Institute for facilitating, supporting, and actively contributing to the underlying project, Practice Monitor for Molecular Diagnostics. Their contributions made this manuscript possible.

## Related Manuscripts Statement

The authors confirm that this manuscript is original and is not currently under consideration, accepted, or published elsewhere. One related publication arising from the same underlying project, *Practice Monitor for Molecular Diagnostics*, is cited in the references (Ref. 22: Pasman R, Jacobs FJC, Eshuis R, Kroon CDd, Sloep M, Werf Jvd, et al. *Integrating Real-World Data and Computational Guidelines: Designing a Dashboard for Nationwide Evaluation of Molecular Diagnostics in Oncology.* IOS Press Ebooks. 2025;15(327):1393-7). This cited work is distinct in scope and content and does not overlap with the data, analyses, or conclusions presented in the current manuscript.

